# Bayesian Multidimensional Nominal Response Model for Observer Study of Radiologists

**DOI:** 10.1101/2022.08.05.22278451

**Authors:** Mizuho Nishio, Daigo Kobayashi, Hidetoshi Matsuo, Yasuyo Urase, Eiko Nishioka, Takamichi Murakami

**Affiliations:** Department of Radiology, Kobe University Graduate School of Medicine, 7-5-2 Kusunoki-cho, Chuo-ku, Kobe 650-0017 JAPAN

**Keywords:** item response theory, nominal response model, probabilistic programming language, chest X-ray

## Abstract

**Purpose:** This study proposes a Bayesian multidimensional nominal response model (MD-NRM) to statistically analyze the nominal response of multiclass classifications.

**Materials and methods:** First, for MD-NRM, we extended the conventional nominal response model to achieve stable convergence of the Bayesian nominal response model and utilized multidimensional ability parameters. We then applied MD-NRM to a 3-class classification problem, where radiologists visually evaluated chest X-ray images and selected their diagnosis from one of the three classes. The classification problem consisted of 150 cases, and each of the six radiologists selected their diagnosis based on a visual evaluation of the images. Consequently, 900 (= 150×6) nominal responses were obtained. In MD-NRM, we assumed that the responses were determined by the softmax function, the ability of radiologists, and the difficulty of images. In addition, we assumed that the multidimensional ability of one radiologist were represented by a 3×3 matrix. The latent parameters of the MD-NRM (ability parameters of radiologists and difficulty parameters of images) were estimated from the 900 responses. To implement Bayesian MD-NRM and estimate the latent parameters, a probabilistic programming language (Stan, version 2.21.0) was used.

**Results:** For all parameters, the Rhat values were less than 1.10. This indicates that the latent parameters of the MD-NRM converged successfully.

**Conclusion:** The results show that it is possible to estimate the latent parameters (ability and difficulty parameters) of the MD-NRM using Stan. Our code for the implementation of the MD-NRM is available as open source.

**Short Abstract:** To statistically analyze the nominal response of multiclass classifications, this study proposes a Bayesian multidimensional nominal response model (MD-NRM). With MD-NRM, it is possible to statistically analyze the nominal response of multiclass classifications obtained by radiologists.

## 1. Introduction

Item response theory (IRT), also known as latent trait theory, is a statistical model/paradigm used for analyzing tests [1–3]. The IRT can be used to evaluate test items and test takers quantitatively and to perform quality assurance of tests. In addition, IRT can be used to equate the test results. Statistical analysis of the IRT was performed using the results of binary classifications (e.g., correct and incorrect answers to test items).

IRT can be implemented using probabilistic programming languages (e.g., WinBUGS, JAGS, and Stan) [4–6]. Extension of IRT is also possible using probabilistic programming languages. For example, the graded response model and nominal response model (NRM) as multiclass extensions of IRT were implemented in Stan [7]. In previous studies, the Bayesian IRT and NRM models were implemented in Stan, and statistical analyses were performed using them [7,8].

In medical diagnosis, the results of various diagnostic procedures are frequently defined as binary classifications: the presence or absence of a disease, the presence or absence of distant metastasis of cancer, the prediction results of cancer death, etc. However, not all medical diagnoses can be expressed as binary classifications, and multiclass classifications are sometimes necessary. For example, the T, N, and M factors of the TNM classification system are expressed as multiclass classifications for many cancer types [9–11]. The statistical analysis of these multiclass classification results requires multiclass analysis (such as NRM).

IRT can be applied to medical diagnosis, by considering (i) the patient as the test item, (ii) the doctor as the test taker, and (iii) the results of the binary classification obtained through medical diagnosis as test results. For example, a previous study used Bayesian IRT to statistically evaluate improvements in radiologists’ diagnostic performance [8]. However, to the best of our knowledge, Bayesian NRM has not been applied to the medical diagnosis of multiclass classifications.

The purpose of this study was to use the Bayesian NRM for multiclass medical diagnosis. Although a Stan implementation of the Bayesian NRM has already been reported [7], it does not work stably [12]. Therefore, for the Bayesian NRM, we extended the conventional NRM in this study. There are two major differences between conventional and our extended NRM. First, while conventional NRM is frequently implemented by extending the 2PL-IRT, our NRM is implemented by extending the 1PL-IRT. We speculate that, because the existing Stan implementation of NRM extends the 2PL-IRT, the existing implementation produced unstable results. The second change in our NRM is to evaluate the ability of test takers using multidimensional parameters rather than a single parameter. Since this study used the results of multiclass classifications, it is natural for test-takers’ abilities to be evaluated based on multidimensional parameters rather than a single parameter. Our Stan implementation of the Bayesian NRM proposed in this study is disclosed as open source through GitHub (https://github.com/jurader/MDNRM).

## 2. Materials and methods

### 2.1. Binary and multiclass classification

Generally, the results of binary classification are summarized by a 2×2 confusion matrix between the ground truth and prediction. This 2×2 confusion matrix is composed of true positive, true negative, false positive, and false negative. Figure 1 shows the 2×2 confusion matrix. In a previous study [8], Bayesian IRT was applied by considering true positive and true negative as “correct” and false positive and false negative as “incorrect”.

**Figure 1.**
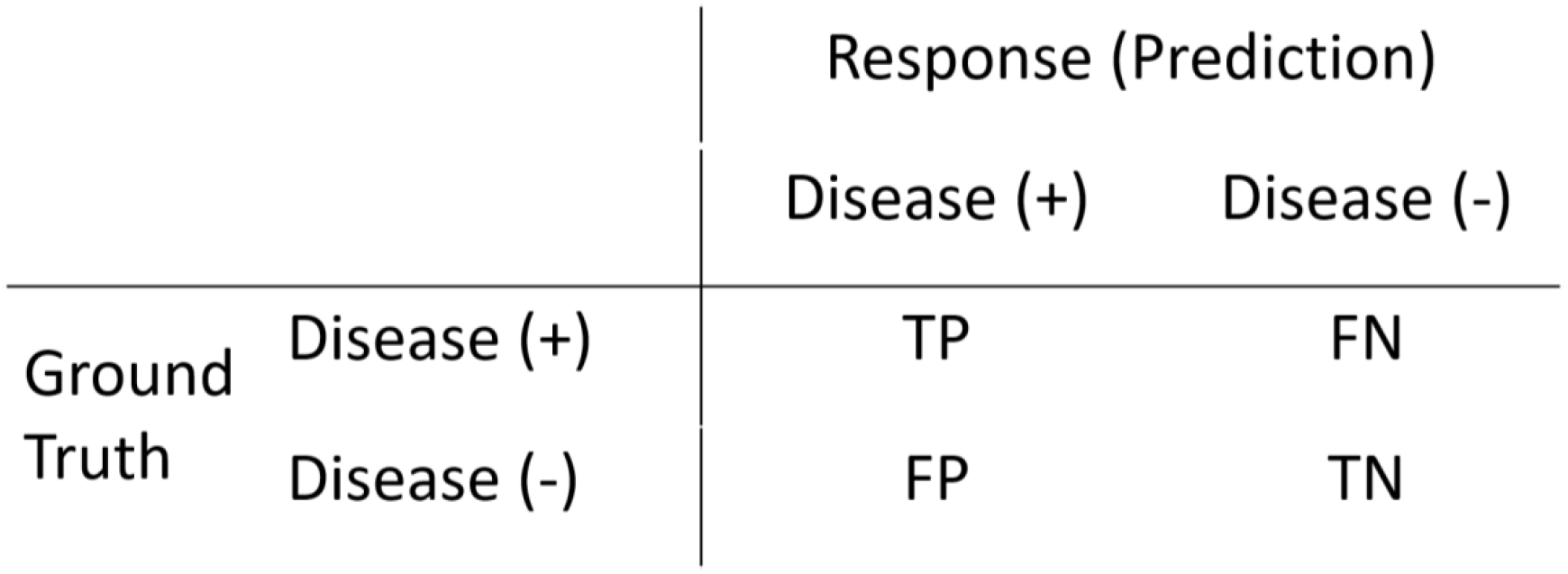
2×2 confusion matrix between ground truth and prediction. Abbreviations: TP, true positive; TN, true negative; FP, false positive; FN, false negative.

In general, the results of multiclass classification are summarized by an N×N confusion matrix. As an example, a 3×3 confusion matrix is shown in Figure 2. Here, it is possible to define the diagonal values of the N×N confusion matrix as “correct” and the other values as “incorrect” and apply Bayesian IRT to these values. However, this method does not allow class-by-class evaluations. To solve this problem, conventional NRM was extended in this study.

**Figure 2.**
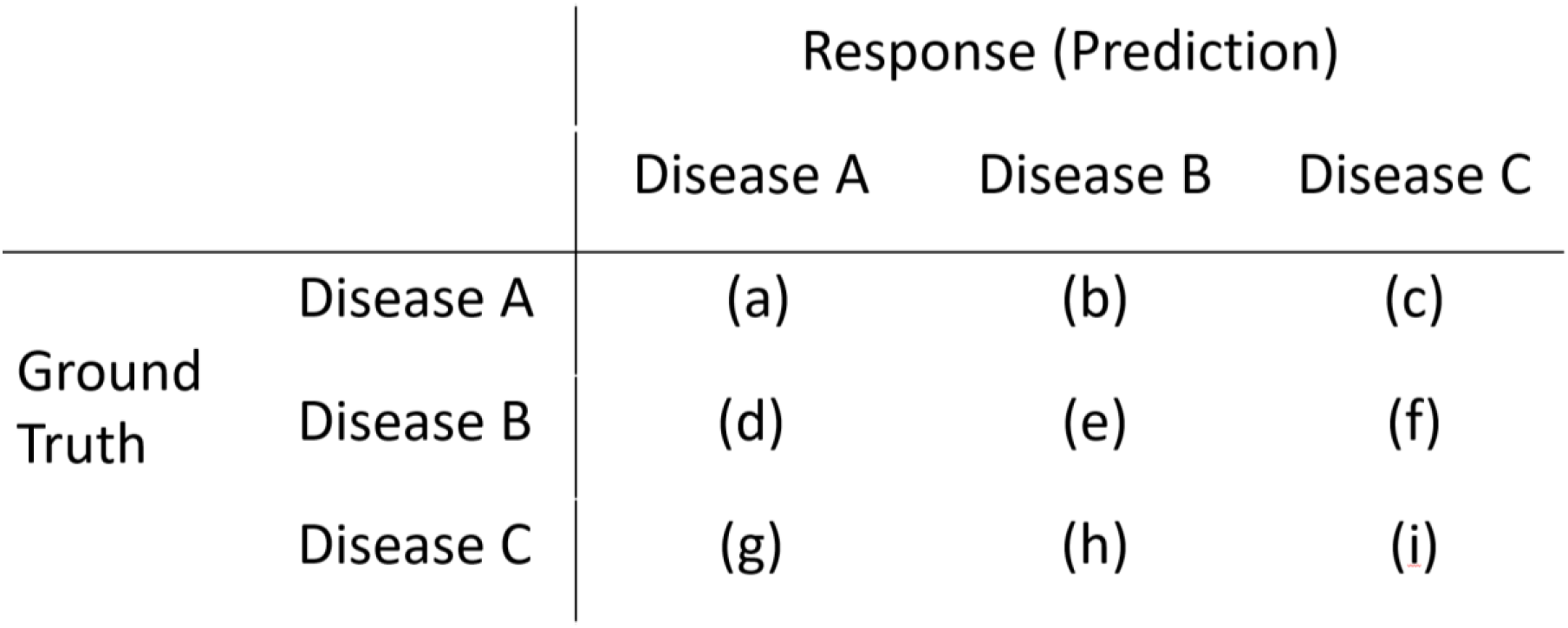
3×3 confusion matrix between ground truth and prediction for 3-class classifications. Note: (a)-(i) represent the frequencies of cells.

### 2.2. IRT and NRM

IRT is a statistical model for analyzing the results of binary classifications. There are several types of IRT models. Here, 1PL-IRT and 2PL-IRT are shown [1]. In addition, conventional NRM, as a multiclass extension of 2PL-IRT, is also described [7]. Finally, an extended NRM is proposed. Hereafter, “case” and “radiologist” are used instead of “test item” and “test taker” of IRT, respectively.

#### 2.2.1. 1PL-IRT

In 1PL-IRT, one parameter (*β*_*i*_) is used to represent case *i*, and another parameter (*θ*_*j*_) is used to represent radiologist *j*. The following equations represent the 1PL-IRT.

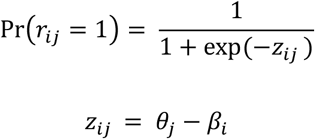

Here,

- *r*_*ij*_ is the response (prediction) of radiologist *j* to case *i*,
- *r*_*ij*_ = 1 means that the response *r*_*ij*_ is correct,
- Pr(*r*_*ij*_ = 1) represents the probability that the response of radiologist *j* to case *i* is correct,
- *β*_*i*_ is the difficulty parameter of case *i*,
- *θ*_*j*_ is the ability parameter of the radiologist *j*.

The equation of 1PL-IRT indicates that for converting logit (*z*_*ij*_) to probability, 1PL-IRT uses the sigmoid function, which is used for logistic regression. In the 1PL-IRT, *θ*_*j*_ *and β*_*i*_ are estimated based on the prediction results of the radiologists.

#### 2.2.2. 2PL-IRT

In the 2PL-IRT, two parameters (*α*_*i*_ and *β*_*i*_) are used to represent case *i*. The following equations represent 2PL-IRT.

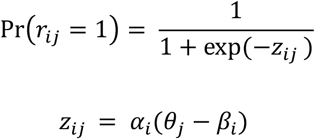

Here,

- *α*_*i*_ and *β*_*i*_ are the discrimination and difficulty parameters of case *i*.

In 2PL-IRT, *θ*_*j*_, *α*_*i*_, and *β*_*i*_ are estimated based on the prediction results of radiologists as in 1PL-IRT.

#### 2.2.3. Conventional NRM

Conventional NRM can be viewed as an extension of 2PL-IRT for multiclass classifications. The following equation represents the conventional NRM.

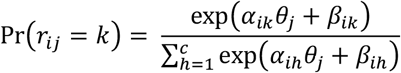

Here,

- Pr(*r*_*ij*_ = *k*) represents the probability that the response of radiologist *j* to case *i* is class *k*,
- The number of classes is *c*,
- *α*_*ik*_ and *β*_*ik*_ are the two parameters of case *i* on class *k*,
- *θ*_*j*_ is the ability parameter of radiologist *j*.

The equation of conventional NRM can be rewritten as follows:

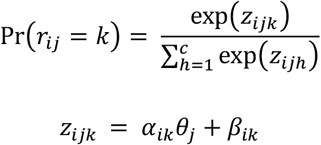

This means that the NRM uses the softmax function to convert logit (*z*_*ijk*_) to probability. Based on the results of the multiclass classification, *α*_*ik*_, *β*_*ik*_, *and θ*_*j*_ are estimated. Although a previous study provided the Stan code for conventional NRM (Bayesian NRM) [7], its calculation results were unstable [12].

#### 2.2.4. Extended NRM (1PL-NRM and multidimensional NRM (MD-NRM))

To stabilize the results of the Bayesian NRM, we extended the conventional NRM. First, the discrimination parameter (*α*) is removed from conventional NRM. In other words, our extended NRM is based on 1PL-IRT. This deletion stabilizes the calculation results of our Bayesian NRM. Hereafter, this extension of NRM is referred to as 1PL-NRM. The following equations represent 1PL-NRM.

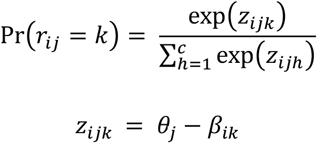

In this equation, *θ*_*j*_ − *β*_*ik*_ is used instead of *θ*_*j*_ + *β*_*ik*_, to strengthen the meaning of the difficulty parameter of *β*_*ik*_.

In both conventional NRM and 1PL-NRM, the probability of *r*_*ij*_ = *k* (the response of radiologist *j* to case *i* is class *k*) depends on only one parameter of the radiologist (*θ*_*j*_). This assumption is unnatural, and the equations of both conventional NRM and 1PL-NRM indicate that the difficulty parameter of the case has a greater influence on which class a radiologist chooses for the diagnosis than their ability parameter.

Because there are *c* classes in multiclass classification, the probability should be dependent on several parameters of the radiologists. To address this problem, we extended 1PL-NRM.

Herein, we propose a multidimensional NRM (MD-NRM) based on 1PL-NRM, which is represented by the following equations:

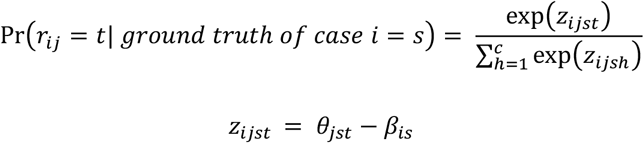

- The number of classes is *c*,
- *s* is the ground truth of case *i*,
- *r*_*ij*_ = *t* means that the response of radiologist *j* to case *i* is *t*,
- *θ*_*jst*_ is the ability parameter of radiologist *j* in class *t* when the ground truth of the case is *s* (*θ*_*j*._ can be represented by a matrix (*c*×*c*)),
- *β*_*is*_ is the difficulty parameter of case *i* on class *s*.

This extended NRM (MD-NRM) means that the multidimensional ability parameters of a radiologist are represented by a matrix, which corresponds to the confusion matrix of multiclass classification.

#### 2.2.5. Relationship between IRT and NRM

The 2PL-IRT can be derived from the conventional NRM as a special case [13]. Here, we use only two classes (*k* or *k*′) for the NRM. Additionally, we consider the conditional probability for a response in class *k* given that the response is one of classes *k* or *k*′. Generally, the conditional probability is given by the following equation:

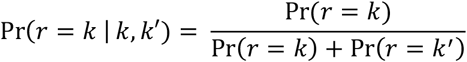

Here, the conventional NRM is used for Pr(*k*) or Pr(*k’*). The conditional probability is represented by the following equation (case *i* and radiologist *j* are omitted for brevity):

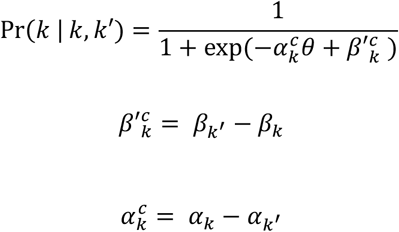

Further, this equation can be converted to the following equations.

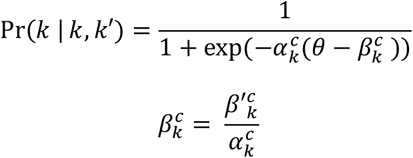

These equations represent the 2PL-IRT. This derivation (conventional NRM to 2PL-IRT) can be applied to our 1PL-NRM; In our 1PL-NRM, the conditional probability is represented by the 1PL-IRT. This is one of the major reasons for using the extended NRM.

### 2.3. Experiments

Our institutional review board approved this retrospective study and waived the requirement for informed consent.

It is assumed that MD-NRM can be applied to data from medical diagnoses of multiclass classification. We applied MD-NRM to the classification results obtained by radiologists in a previous study [14]. In the previous study, there were three classes of medical diagnosis: novel coronavirus pneumonia (COVID), non-novel-coronavirus pneumonia (PNEUMONIA), and normal (NORMAL). Therefore, the classification results were summarized as a 3×3 confusion matrix. In total, 150 cases (50 COVID, 50 PNEUMONIA, and 50 NORMAL) were reported in the previous study. From the three classes, six radiologists determined their diagnoses based on visual evaluation of chest X-ray images. Therefore, 900 (150×6) nominal responses were obtained. The Supplementary material shows the ground truth of the case and the classification results of the six radiologists.

In this study, MD-NRM was applied to multiclass classification results to analyze the ability of six radiologists for the three classes. For all ability and difficulty parameters, a normal distribution with mean = 0 and standard deviation = 2 was used as the prior distribution. The following parameters were used for sampling in Stan: chains=8, iter=8000, warmup=4000, thin=1, adapt_delta = 0.9, and max_treedepth = 15. The convergence check of the MD-NRM was performed by evaluating the Rhat values of all parameters [7,8,15]. Since we focused on the ability parameters, the estimation results of the difficulty parameters were omitted in this study. We used the following software to implement MD-NRM: R, version 4.1.1; Stan, version 2.21.0; rstan, version 2.21.2; shinystan, version 2.5.0; and tidybayes, version 3.0.1.

## 3. Results

Figure 3 shows the convergence check of the Stan results for all the parameters. The Rhat values for all parameters (difficulty and ability parameters) were less than 1.10, which means that the MD-NRM converged successfully [7,8,15]. Thus, 1PL-NRM is useful for stabilizing Bayesian NRM.

**Figure 3.**
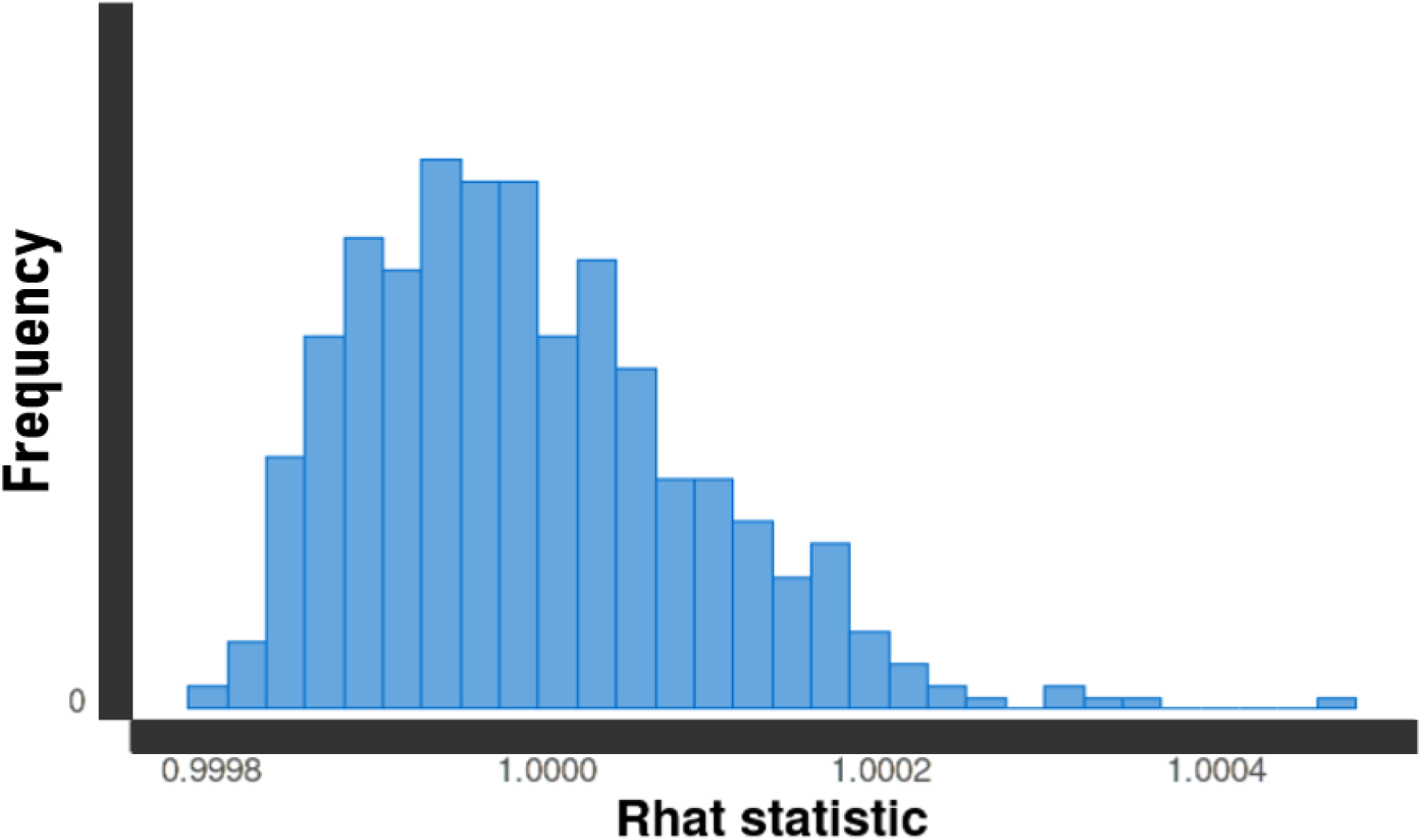
Evaluation of convergence in MD-NRM. Note: For all parameters, their Rhat values are less than 1.10.

Table 1 shows the estimation results of the MD-NRM for the ability parameters of Radiologist 1. In the MD-NRM, radiologist ability is represented in a 3×3 matrix, so one radiologist has nine ability parameters. In Table 1, if *θ*_111_, *θ*_122_, and *θ*_133_ are high, the diagnostic performance of radiologist 1 is high for NORMAL, PNEUMONIA, and COVID.

**Table 1.** Result of MD-NRM for ability parameters of radiologist 1. Note: 2.50%, 25%, 50%, 75%, and 97.5% are percentile values of the estimated parameters. For example, the pair of −0.137 at 2.50% and 4.665 at 97.5% represents a 95% credible interval of *θ*_111_.

| Parameters | 2.50% | 25% | 50% (median) | 75% | 97.50% |
| --- | --- | --- | --- | --- | --- |
| $\theta_{111}$ | -0.137 | 1.46 | 2.288 | 3.117 | 4.665 |
| $\theta_{112}$ | -2.835 | -1.242 | -0.403 | 0.452 | 2.035 |
| $\theta_{113}$ | -4.44 | -2.763 | -1.896 | -1.009 | 0.655 |
| $\theta_{121}$ | -4.659 | -2.987 | -2.137 | -1.262 | 0.372 |
| $\theta_{122}$ | -0.796 | 0.762 | 1.589 | 2.418 | 3.969 |
| $\theta_{123}$ | -1.845 | -0.268 | 0.556 | 1.387 | 2.959 |
| $\theta_{131}$ | -3.652 | -2.077 | -1.249 | -0.422 | 1.155 |
| $\theta_{132}$ | -2.572 | -1.049 | -0.233 | 0.593 | 2.122 |
| $\theta_{133}$ | -0.831 | 0.682 | 1.491 | 2.297 | 3.824 |

Figure 4 shows a 3 × 3 ability matrix created from the median values of the ability parameters of radiologist 1 in Table 1. In Figure 4, if the diagonal values of the ability matrix are high, the diagnostic ability of radiologist 1 is also high. In contrast, if the non-diagonal values of the ability matrix are low, the diagnostic ability is high.

**Figure 4.**
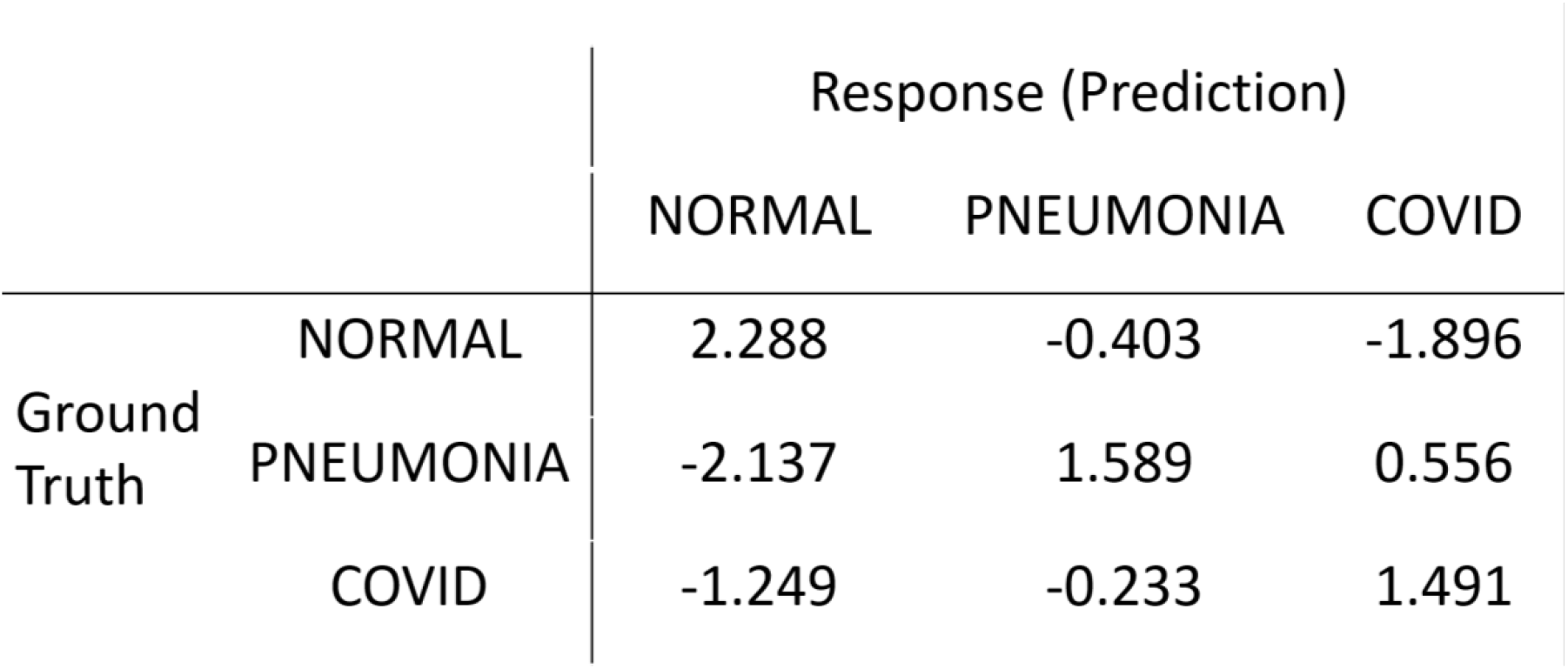
Ability matrix of radiologist 1. Note: This matrix was created from the median values of the ability parameters of radiologist 1 in Table 1.

Figure 5 (A)–(C) show the summary of *θ*_*i*11_ (*i* = 1, 2, …, 6), *θ*_*i*22_ (*i* = 1, 2, …, 6), and *θ*_*i*33_ (*i* = 1, 2, …, 6) for radiologists 1–6 for NORMAL, PNEUMONIA, and COVID, respectively. The figures in the Supplementary Materials show all the ability parameters of radiologists 1–6.

**Figure 5.**
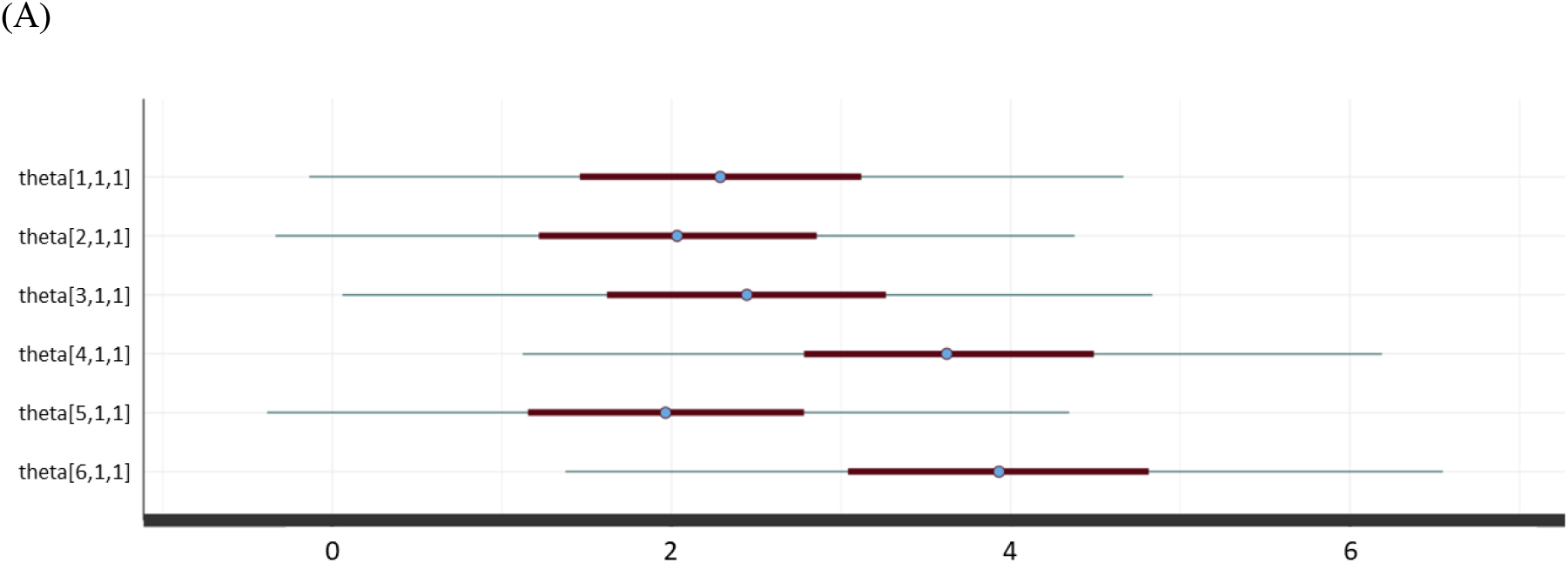
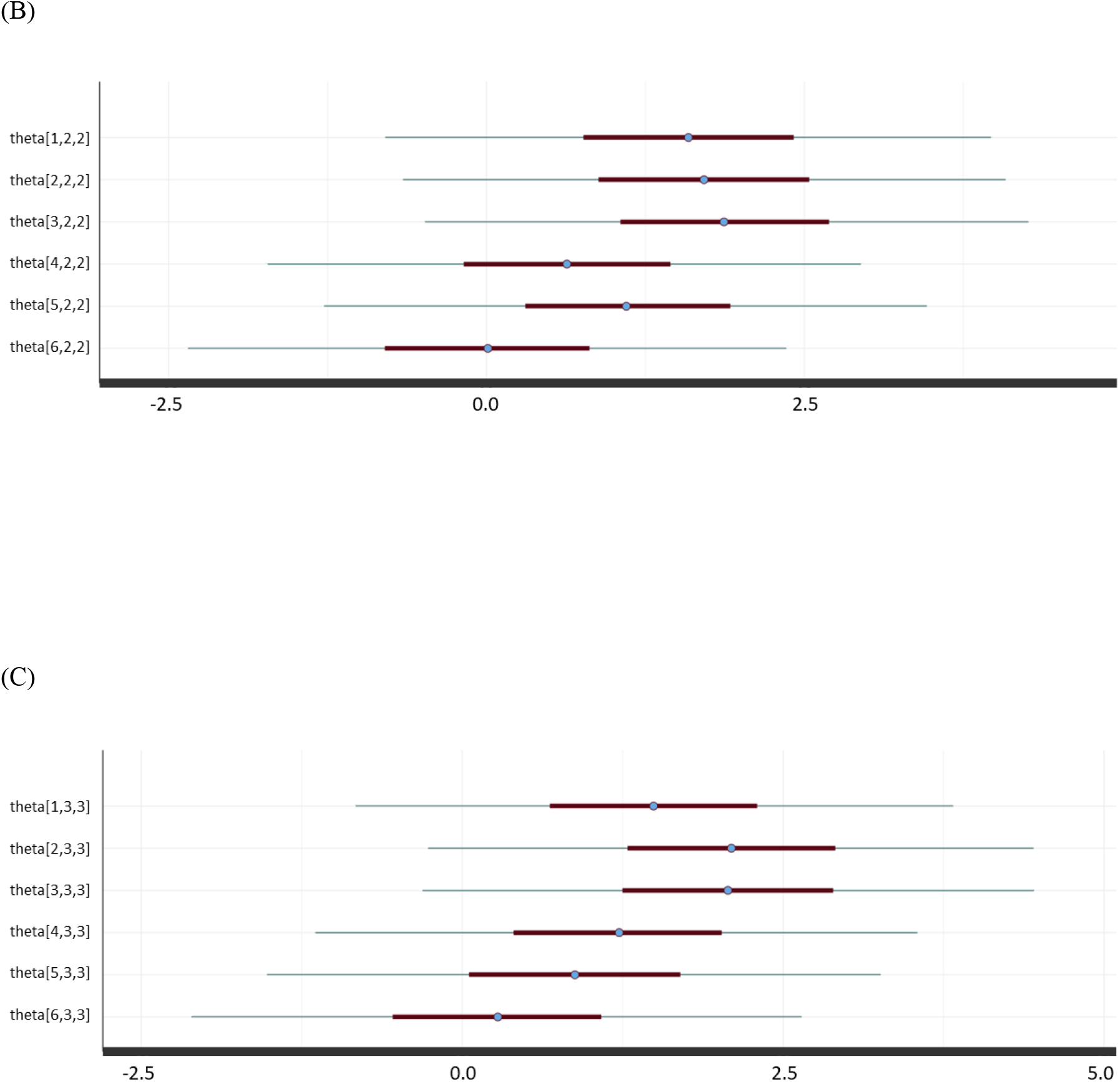
Summary of ability parameters of six radiologist. (A) *θ*_*i*11_ (*i* = 1, 2, …, 6) for NORMAL, (B) *θ*_*i*22_ (*i* = 1, 2, …, 6) for PNEUMONIA, (C) *θ*_*i*33_ (*i* = 1, 2, …, 6) for COVID. Note: In (A)–(C), “theta[i,j,k]” represents *θ*_*ijk*_. For example, “theta[1,1,1]” represents *θ*_111_. Circles, thick bars, and thin bars represent the median values, 50% credible interval (interquartile range), and 95% credible interval, respectively.

Our Stan and R codes of the MD-NRM are shown in the Supplementary materials. In addition, these codes were disclosed as open source through GitHub (https://github.com/jurader/MDNRM).

## 4. Discussion

Using the MD-NRM, stable convergence in the Bayesian NRM calculation was achieved with our Stan code. This is a significant improvement because stable convergence cannot be obtained with the conventional NRM Stan code [7]. In addition, compared to the conventional NRM, MD-NRM makes it possible to determine the multidimensional ability parameters of a doctor. Furthermore, this extension of multidimensional parameters allows MD-NRM to provide a more detailed evaluation of a doctor’s ability rather than a single parameter.

In IRT and NRM, parameters (ability and difficulty) are evaluated in the latent space [1,8]. Because of this characteristic, IRT is known as the latent trait theory. Logistic regression is frequently used to analyze the results of medical diagnoses [16]. In logistic regression, the effect of covariates on the latent space is statistically evaluated. Therefore, IRT, NRM, and logistic regression share similar functionalities in the evaluation of the parameters.

In receiver operating characteristic (ROC) analysis, it is common to assign a multilevel score for each case to statistically analyze the results of binary classifications [17,18]. For example, a radiologist’s ability to differentiate between benign and malignant tumors is frequently analyzed by assigning a score to each case on a 5-point scale [19]. However, there is no commonly used method of ROC analysis for multiclass classification results. Instead, for the analysis of multiclass classifications, it is relatively frequent to perform multiple ROC analyses of binary classifications in a one-vs-rest fashion [20]. On the other hand, MD-NRM can analyze the results of multiclass classifications by a single analysis, which is a major difference between MD-NRM and ROC analysis. In addition, MD-NRM does not require the multilevel scoring (please see the data of 150 cases in the Supplementary material), which is another major difference from ROC analysis.

In a previous study [8], it was possible to introduce covariates into MD-NRM. This extension makes it possible to quantitatively evaluate the effect of covariates using MD-NRM; for example, the improvement in diagnostic performance with various experiences of doctors or with different diagnostic modalities/examinations.

This study had several limitations. First, the data from 150 patients were used. It has not been evaluated whether stable convergence will be obtained in MD-NRM with smaller data. Second, although an extension of the MD-NRM (MD-NRM with covariates) is proposed in this paper, no actual experiments have been conducted. We expect that the extension of MD-NRM will be investigated in the future based on our open-source code.

In conclusion, the MD-NRM was proposed as a statistical analysis method for multiclass classification. The MD-NRM achieved successful convergence of parameter estimation in the Bayesian NRM. In addition, using the MD-NRM, a more detailed evaluation of the doctor’s ability was possible by using multidimensional ability parameters.

## Data Availability

All data produced in the present study are available upon reasonable request to the authors

https://github.com/jurader/MDNRM

## Acknowledgment

We thank Yoshiaki Watanabe (Nishinomiya Watanabe Hospital) for his assistance.

## Source code

In addition to the Supplementary material, the source code of our MD-NRM is available from the following URL of GitHub.

https://github.com/jurader/MDNRM

## Supplementary material

- Data of ground truth and radiologists’ prediction for 150 patients
- Ability parameters of radiologists 1–6.
- R code
- Stan code

| GroundTruth | Rad1 | Rad2 | Rad3 | Rad4 | Rad5 | Rad6 |
| --- | --- | --- | --- | --- | --- | --- |
| 1 | 1 | 0 | 1 | 1 | 1 | 1 |
| 1 | 1 | 1 | 1 | 1 | 1 | 0 |
| 0 | 1 | 0 | 0 | 0 | 0 | 0 |
| 1 | 1 | 1 | 2 | 2 | 2 | 2 |
| 1 | 1 | 1 | 1 | 1 | 2 | 2 |
| 1 | 2 | 1 | 1 | 1 | 1 | 0 |
| 1 | 2 | 0 | 1 | 0 | 0 | 0 |
| 0 | 2 | 2 | 0 | 1 | 0 | 0 |
| 2 | 2 | 1 | 2 | 2 | 1 | 1 |
| 2 | 0 | 0 | 0 | 0 | 0 | 0 |
| 2 | 2 | 2 | 2 | 2 | 2 | 0 |
| 0 | 0 | 0 | 2 | 0 | 2 | 0 |
| 1 | 2 | 2 | 1 | 2 | 2 | 0 |
| 0 | 0 | 0 | 0 | 0 | 0 | 0 |
| 0 | 0 | 0 | 0 | 0 | 0 | 0 |
| 0 | 1 | 0 | 1 | 1 | 0 | 0 |
| 1 | 2 | 0 | 1 | 0 | 2 | 0 |
| 1 | 2 | 2 | 2 | 2 | 2 | 2 |
| 1 | 2 | 1 | 1 | 1 | 1 | 0 |
| 1 | 2 | 2 | 2 | 0 | 2 | 2 |
| 1 | 2 | 1 | 2 | 2 | 1 | 2 |
| 1 | 1 | 1 | 2 | 1 | 1 | 1 |
| 1 | 1 | 1 | 1 | 1 | 1 | 1 |
| 0 | 0 | 0 | 0 | 0 | 2 | 0 |
| 2 | 0 | 2 | 2 | 0 | 2 | 0 |
| 2 | 1 | 2 | 2 | 0 | 0 | 0 |
| 2 | 0 | 0 | 0 | 0 | 2 | 0 |
| 1 | 2 | 1 | 1 | 2 | 1 | 2 |
| 1 | 1 | 2 | 1 | 1 | 2 | 1 |
| 0 | 1 | 0 | 0 | 0 | 0 | 0 |
| 1 | 1 | 1 | 1 | 1 | 2 | 1 |
| 0 | 0 | 0 | 0 | 0 | 0 | 0 |
| 0 | 0 | 0 | 0 | 0 | 1 | 0 |
| 2 | 1 | 2 | 2 | 0 | 1 | 1 |
| 0 | 0 | 0 | 0 | 0 | 0 | 0 |
| 1 | 1 | 2 | 1 | 2 | 1 | 2 |
| 0 | 0 | 1 | 0 | 0 | 1 | 0 |
| 0 | 2 | 0 | 2 | 0 | 2 | 0 |
| 0 | 0 | 0 | 0 | 0 | 0 | 0 |
| 1 | 2 | 2 | 1 | 0 | 2 | 0 |
| 1 | 0 | 0 | 0 | 0 | 0 | 0 |
| 1 | 2 | 2 | 1 | 2 | 1 | 0 |
| 0 | 1 | 1 | 0 | 0 | 1 | 0 |
| 1 | 1 | 1 | 1 | 1 | 1 | 1 |
| 0 | 0 | 0 | 0 | 0 | 0 | 0 |
| 2 | 2 | 2 | 2 | 2 | 2 | 2 |
| 0 | 0 | 0 | 0 | 0 | 2 | 0 |
| 0 | 0 | 0 | 0 | 0 | 0 | 0 |
| 0 | 0 | 0 | 1 | 0 | 0 | 0 |
| 1 | 1 | 1 | 1 | 1 | 2 | 0 |
| 1 | 1 | 1 | 2 | 1 | 1 | 1 |
| 0 | 0 | 0 | 0 | 0 | 0 | 0 |
| 2 | 2 | 2 | 2 | 0 | 2 | 0 |
| 2 | 2 | 2 | 2 | 2 | 1 | 2 |
| 1 | 1 | 1 | 1 | 2 | 2 | 2 |
| 2 | 2 | 1 | 2 | 2 | 2 | 2 |
| 2 | 1 | 2 | 1 | 2 | 1 | 2 |
| 0 | 0 | 0 | 0 | 0 | 0 | 0 |
| 2 | 2 | 0 | 2 | 0 | 0 | 0 |
| 2 | 2 | 2 | 2 | 0 | 0 | 0 |
| 2 | 2 | 2 | 2 | 2 | 2 | 0 |
| 0 | 1 | 0 | 2 | 0 | 0 | 0 |
| 0 | 0 | 1 | 1 | 0 | 0 | 0 |
| 0 | 0 | 2 | 0 | 0 | 0 | 1 |
| 1 | 0 | 2 | 2 | 2 | 2 | 1 |
| 1 | 2 | 1 | 1 | 1 | 1 | 1 |
| 2 | 0 | 2 | 2 | 0 | 2 | 0 |
| 1 | 2 | 1 | 1 | 2 | 2 | 1 |
| 1 | 1 | 1 | 1 | 1 | 1 | 1 |
| 0 | 0 | 0 | 0 | 0 | 0 | 0 |
| 0 | 1 | 0 | 0 | 0 | 1 | 0 |
| 2 | 2 | 2 | 2 | 1 | 2 | 1 |
| 2 | 2 | 2 | 2 | 2 | 2 | 2 |
| 1 | 1 | 1 | 1 | 1 | 1 | 2 |
| 0 | 0 | 0 | 0 | 0 | 0 | 0 |
| 1 | 1 | 1 | 2 | 2 | 1 | 1 |
| 0 | 0 | 0 | 0 | 0 | 0 | 0 |
| 0 | 0 | 2 | 1 | 0 | 0 | 0 |
| 2 | 1 | 1 | 1 | 2 | 2 | 2 |
| 0 | 0 | 0 | 0 | 0 | 0 | 0 |
| 0 | 0 | 0 | 0 | 0 | 1 | 0 |
| 1 | 2 | 1 | 1 | 0 | 1 | 0 |
| 2 | 0 | 2 | 2 | 0 | 2 | 0 |
| 0 | 0 | 0 | 0 | 0 | 0 | 0 |
| 0 | 0 | 1 | 0 | 0 | 0 | 0 |
| 1 | 1 | 1 | 1 | 1 | 1 | 2 |
| 2 | 2 | 2 | 2 | 0 | 2 | 0 |
| 2 | 2 | 2 | 2 | 0 | 2 | 0 |
| 0 | 0 | 2 | 0 | 0 | 0 | 0 |
| 2 | 2 | 2 | 1 | 2 | 2 | 0 |
| 0 | 0 | 1 | 0 | 0 | 0 | 0 |
| 1 | 1 | 1 | 1 | 1 | 2 | 0 |
| 2 | 2 | 2 | 2 | 2 | 2 | 2 |
| 0 | 0 | 0 | 0 | 0 | 0 | 0 |
| 2 | 1 | 2 | 2 | 0 | 0 | 0 |
| 2 | 1 | 2 | 2 | 2 | 1 | 0 |
| 0 | 0 | 0 | 0 | 0 | 0 | 0 |
| 2 | 2 | 1 | 2 | 2 | 1 | 2 |
| 0 | 0 | 0 | 0 | 0 | 0 | 0 |
| 1 | 1 | 2 | 1 | 2 | 2 | 2 |
| 1 | 1 | 2 | 1 | 1 | 1 | 2 |
| 1 | 0 | 0 | 0 | 0 | 2 | 0 |
| 2 | 0 | 2 | 2 | 0 | 0 | 0 |
| 1 | 1 | 1 | 1 | 1 | 1 | 0 |
| 1 | 1 | 1 | 0 | 2 | 2 | 0 |
| 0 | 0 | 0 | 0 | 0 | 1 | 0 |
| 1 | 2 | 1 | 2 | 2 | 2 | 2 |
| 2 | 2 | 2 | 0 | 0 | 2 | 0 |
| 2 | 2 | 2 | 2 | 2 | 2 | 2 |
| 2 | 2 | 1 | 2 | 2 | 1 | 1 |
| 2 | 1 | 1 | 1 | 0 | 0 | 0 |
| 1 | 2 | 1 | 1 | 2 | 1 | 1 |
| 0 | 0 | 0 | 0 | 0 | 0 | 0 |
| 0 | 0 | 0 | 0 | 0 | 0 | 0 |
| 0 | 0 | 2 | 0 | 0 | 0 | 0 |
| 1 | 1 | 1 | 1 | 1 | 2 | 1 |
| 2 | 1 | 2 | 1 | 2 | 1 | 1 |
| 0 | 1 | 0 | 0 | 0 | 1 | 0 |
| 1 | 1 | 1 | 2 | 0 | 1 | 1 |
| 0 | 0 | 0 | 0 | 0 | 0 | 0 |
| 0 | 0 | 0 | 0 | 0 | 0 | 0 |
| 1 | 1 | 1 | 1 | 1 | 1 | 1 |
| 2 | 2 | 2 | 2 | 0 | 1 | 0 |
| 2 | 1 | 1 | 1 | 2 | 1 | 2 |
| 2 | 2 | 2 | 2 | 2 | 2 | 2 |
| 0 | 0 | 0 | 0 | 0 | 0 | 0 |
| 1 | 1 | 1 | 0 | 0 | 0 | 0 |
| 2 | 2 | 2 | 2 | 2 | 1 | 1 |
| 2 | 2 | 2 | 2 | 2 | 2 | 0 |
| 2 | 2 | 2 | 1 | 2 | 2 | 2 |
| 2 | 2 | 2 | 2 | 2 | 2 | 2 |
| 2 | 1 | 2 | 2 | 0 | 0 | 0 |
| 0 | 0 | 0 | 0 | 0 | 0 | 0 |
| 2 | 1 | 1 | 1 | 2 | 1 | 2 |
| 0 | 0 | 0 | 0 | 0 | 0 | 0 |
| 2 | 2 | 2 | 2 | 2 | 2 | 2 |
| 2 | 2 | 2 | 2 | 2 | 0 | 0 |
| 1 | 1 | 1 | 1 | 1 | 1 | 0 |
| 0 | 0 | 0 | 0 | 0 | 0 | 0 |
| 1 | 1 | 1 | 1 | 0 | 0 | 0 |
| 2 | 2 | 1 | 1 | 1 | 1 | 2 |
| 2 | 2 | 2 | 2 | 2 | 2 | 1 |
| 1 | 1 | 1 | 1 | 1 | 1 | 1 |
| 0 | 0 | 0 | 0 | 0 | 0 | 0 |
| 2 | 0 | 2 | 2 | 0 | 0 | 0 |
| 1 | 1 | 1 | 1 | 2 | 2 | 2 |
| 1 | 1 | 1 | 1 | 1 | 1 | 1 |
| 2 | 2 | 2 | 1 | 2 | 2 | 2 |
| 2 | 2 | 2 | 2 | 0 | 1 | 2 |
| 2 | 2 | 1 | 2 | 1 | 1 | 1 |

**Figure S1.**
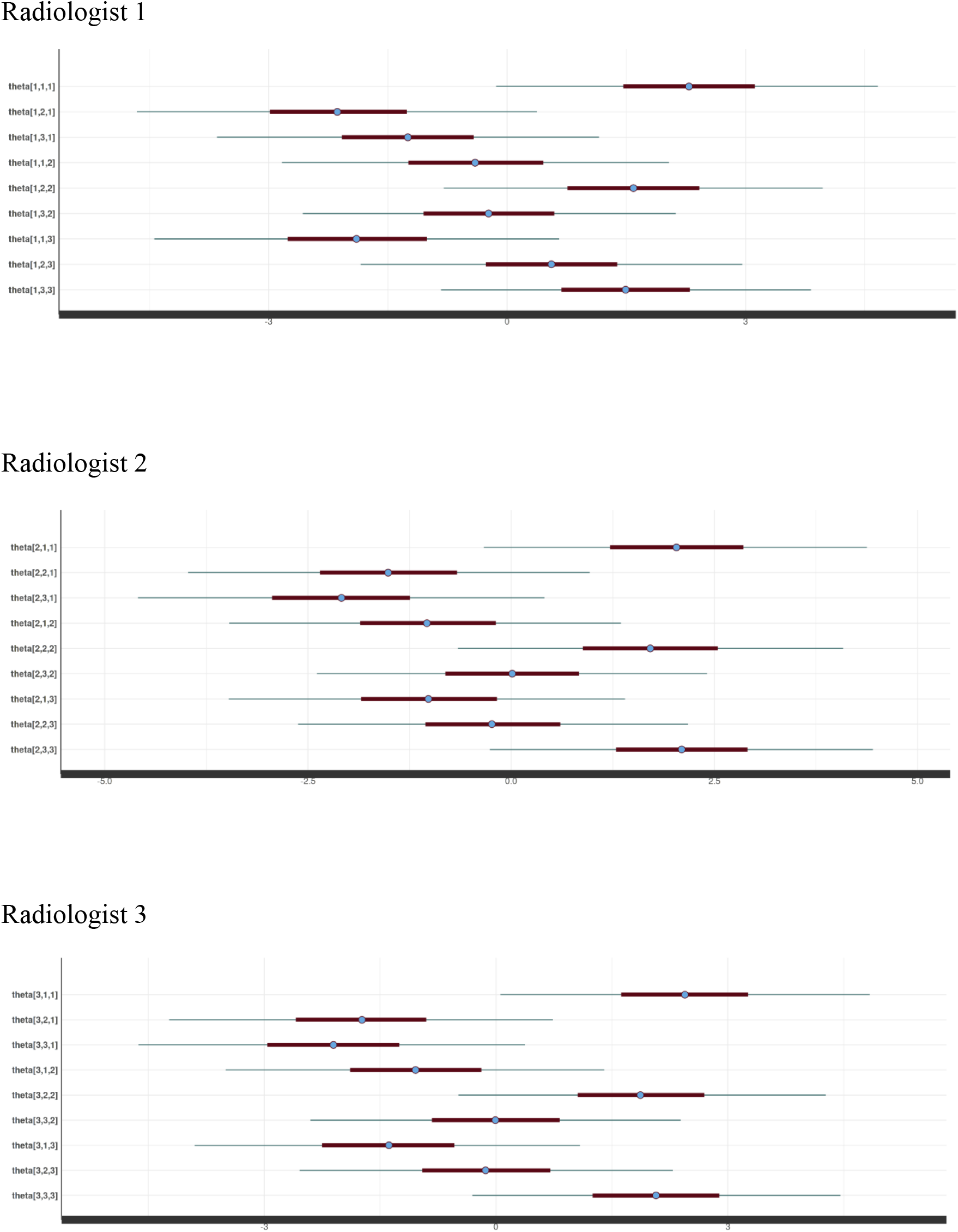
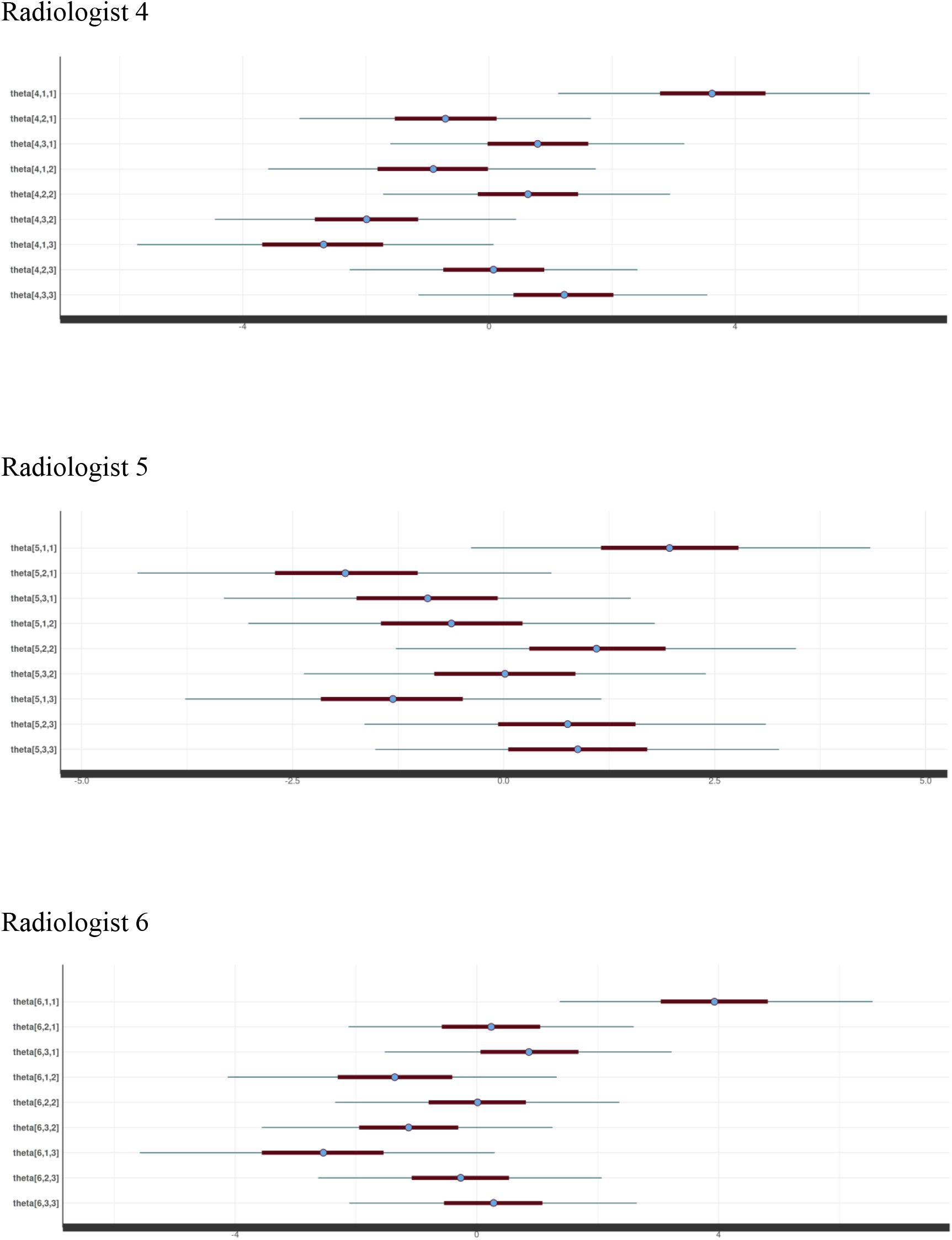
Ability parameters of radiologists 1–6.

~~~
#csv_path = “./my_stan_code/ground_truth_and_results.csv”
csv_path = “./ground_truth_and_results.csv”
#path_stan = “./my_stan_code/my_NRM2.stan”
path_stan = “./my_stan_code/MDNRM.stan”
MY_SEED <- 1234
library(rstan)
library(dplyr)
library(tidybayes)
########################
########################
########################
resp <- read.csv(csv_path)
resp <- resp+1
# After the above line
# 1 -> normal
# 2 -> non-COVID19 pneumonia
# 3 -> COVID19 pneumonia
types <- c(resp$GT)
resp <- resp[,2:7]
resp <- t(resp)
N <- nrow(resp)
T <- ncol(resp)
print(csv_path)
print(“number of radiologists:”)
print(N)
print(“number of cases (problems):”)
print(T)
head(resp)
head(types)
data_nrm <- list(n_doctor=N, n_case=T, response=resp, K=3, types=types)
print(“starting stan model …”)
print(path_stan)
rstan_options(auto_write=TRUE)
options(mc.cores=parallel::detectCores())
model.NRM <- stan_model(path_stan)
fit.mcmc_nrm <- sampling(model.NRM, data=data_nrm, chains=8, iter=8000,
warmup=4000, thin=1, seed=MY_SEED, control = list(adapt_delta = 0.9,
max_treedepth = 15))
print(fit.mcmc_nrm)
fit.mcmc_nrm %>% spread_draws(beta[n_case, K]) %>% median_qi(.width = c(.95))
-> beta
print(“***** beta *****”)
#print(beta)
#head(beta, 10)
print(beta, n=450)
fit.mcmc_nrm %>% spread_draws(theta[n_doctor, K1, K2]) %>% median_qi(.width
= c(.95)) -> theta
print(“***** theta *****”)
#print(theta)
#head(theta, 10)
print(theta, n=100)
# check Rhat values
rs = summary(fit.mcmc_nrm)$summary[,”Rhat”]
print(“***** Rhat *****”)
print(summary(rs))
print(all(rs < 1.10, na.rm=T))
#For GUI
#library(shinystan)
#launch_shinystan(fit.mcmc_nrm)
data{
  int<upper=3> K; // number of clesses of responses
  int<lower=1> n_doctor; // number of doctors
  int<lower=1> n_case; // number of cases (problems)
  int<lower=1,upper=K> response[n_doctor,n_case]; //array of responses
  int<lower=1,upper=3> types[n_case]; //type of cases (problems)
}
parameters {
  vector[K] beta[n_case]; // difficulty
  matrix[K,K] theta[n_doctor]; // ability
}
model{
  int t;
  vector[K] target_theta;
//prior
for (i in 1:n_doctor){
  for (j in 1:K){
    for (k in 1:K){
      theta[i,j,k] ∼ normal(0,2);
    }
  }
}
for (i in 1:n_case){
  beta[i] ∼ normal(0,2);
}
// likelihood
for (i in 1:n_doctor){
  for (j in 1:n_case){
    t = types[j];
    target_theta = to_vector(theta[i,t]);
    response[i,j] ∼ categorical_logit(-beta[j] + target_theta);
  }
 }
}
~~~

## Notes

**<u>Funding information</u>** This work was supported by JSPS KAKENHI (Grant Numbers: JP19K17232 and 22K07665).

**<u>Conflict of Interest</u>** There are no conflicts of interest to declare.

### Competing Interest Statement

The authors have declared no competing interest.

### Funding Statement

This work was supported by JSPS KAKENHI (Grant Numbers: JP19K17232 and 22K07665).

### Author Declarations

IRB of KOBE University Hospital gave ethical approval for this work.

## References

1. Cappelleri JC, Jason Lundy J, Hays RD. Overview of classical test theory and item response theory for the quantitative assessment of items in developing patient-reported outcomes measures. Clin Ther. Clin Ther; 2014;36:648–62.

2. Embretson SE, Reise SP. Item response theory for psychologists. Item Response Theory for Psychologists. Taylor and Francis; 2000.

3. Hays RD, Morales LS, Reise SP. Item response theory and health outcomes measurement in the 21st century. Med Care. 2000;38:II28–42.

4. Carpenter B, Gelman A, Hoffman MD, Lee D, Goodrich B, Betancourt M, et al. Stan: A Probabilistic Programming Language. Journal of Statistical Software. 2017;76:1–32.

5. Lunn DJ, Thomas A, Best N, Spiegelhalter D. WinBUGS - A Bayesian modelling framework: Concepts, structure, and extensibility. Statistics and Computing. 2000;10:325–37.

6. Depaoli S, Clifton JP, Cobb PR. Just Another Gibbs Sampler (JAGS). Journal of Educational and Behavioral Statistics. 2016;41:628–49.

7. Luo Y, Jiao H. Using the Stan Program for Bayesian Item Response Theory: Educational and Psychological Measurement. SAGE PublicationsSage CA: Los Angeles, CA; 2017;78:384–408.

8. Nishio M, Akasaka T, Sakamoto R, Togashi K. Bayesian Statistical Model of Item Response Theory in Observer Studies of Radiologists. Academic Radiology. Elsevier; 2020;27:e45–54.

9. Lim W, Ridge CA, Nicholson AG, Mirsadraee S. The 8th lung cancer TNM classification and clinical staging system: review of the changes and clinical implications. Quantitative Imaging in Medicine and Surgery. AME Publications; 2018;8:709–18.

10. Kalli S, Semine A, Cohen S, Naber SP, Makim SS, Bahl M. American joint committee on cancer’s staging system for breast cancer, eighth edition: What the radiologist needs to know. Radiographics. Radiological Society of North America Inc.; 2018;38:1921–33.

11. Allen PJ, Kuk D, Castillo CF del, Basturk O, Wolfgang CL, Cameron JL, et al. Multiinstitutional Validation Study of the American Joint Commission on Cancer (8th Edition) Changes for T and N Staging in Patients With Pancreatic Adenocarcinoma. Ann Surg. Ann Surg; 2017;265:185–91.

12. Non convergence issue on polytomous IRT model - Modeling - The Stan Forums [Internet]. [cited 2022 Jul 12]. Available from: https://discourse.mc-stan.org/t/non-convergence-issue-on-polytomous-irt-model/12576

13. Thissen D, Cai L, Bock RD. The Nominal Categories Item Response Model. Handbook of Polytomous Item Response Theory Models. Routledge Handbooks Online; 2010.

14. Nishio M, Kobayashi D, Nishioka E, Matsuo H, Urase Y, Onoue K, et al. Deep learning model for the automatic classification of COVID-19 pneumonia, non-COVID-19 pneumonia, and the healthy: a multi-center retrospective study. Sci Rep. Sci Rep; 2022;12:8214.

15. Gelman A, Carlin JB, Stern HS, Dunson DB, Vehtari A, Rubin DB. Bayesian Data Analysis. Bayesian Data Analysis. Chapman and Hall/CRC; 2013.

16. Kleinbaum DG, Klein M. Logistic Regression. New York, NY: Springer; 2010.

17. Obuchowski NA. Receiver operating characteristic curves and their use in radiology. Radiology. Radiology; 2003;229:3–8.

18. Zweig MH, Campbell G. Receiver-operating characteristic (ROC) plots: a fundamental evaluation tool in clinical medicine. Clinical Chemistry. Oxford Academic; 1993;39:561–77.

19. Park SH, Goo JM, Jo CH. Receiver Operating Characteristic (ROC) Curve: Practical Review for Radiologists. Korean Journal of Radiology. Korean Society of Radiology; 2004;5:18.

20. Receiver Operating Characteristic (ROC) — scikit-learn 1.1.1 documentation [Internet]. [cited 2022 Jul 12]. Available from: https://scikit-learn.org/stable/auto_examples/model_selection/plot_roc.html#plot-roc-curves-for-the-multiclass-problem

